# Evaluation of the CD4+ T cell response to SARS-CoV-2 infection and cross reactivity to beta variant in children of all ages

**DOI:** 10.1101/2022.01.27.22269976

**Authors:** Kevin Paul, Freya Sibbertsen, Daniela Weiskopf, Marc Lütgehetmann, Madalena Barroso, Marta K. Danecka, Laura Glau, Laura Hecher, Katharina Hermann, Aloisa Kohl, Jun Oh, Julian Schulze zur Wiesch, Alessandro Sette, Eva Tolosa, Eik Vettorazzi, Mathias Woidy, Antonia Zapf, Dimitra E. Zazara, Thomas S. Mir, Ania C. Muntau, Søren W. Gersting, Gábor A. Dunay

**Author notes:** Correspondence to Gábor A. Dunay, University Children’s Research, UCR@Kinder-UKE, University, Medical Center Hamburg-Eppendorf, Martinistr. 52, 20251 Hamburg, Germany.

## Abstract

SARS-CoV-2 is still a major burden for global health despite effective vaccines. With the reduction of social distancing measures, infection rates are increasing in children, while data on the pediatric immune response to SARS-CoV-2 infection is still lacking. Although the typical disease course in children has been mild, emerging variants may present new challenges in this age group.

Peripheral blood mononuclear cells (PBMC) from 51 convalescent children, 24 seronegative siblings from early 2020, and 51 unexposed controls were stimulated with SARS-CoV-2-derived peptide MegaPools from the ancestral and beta variants. Flow cytometric determination of activation-induced markers and secreted cytokines were used to quantify the CD4+ T cell response.

The average time after infection was over 80 days. CD4+ T cell responses were detected in 61% of convalescent children and were markedly reduced in preschool children. Cross-reactive T cells for the SARS-CoV-2 beta variant were identified in 45% of cases after infection with an ancestral SARS-CoV-2 variant. The CD4+ T cell response was accompanied most predominantly by IFN-*γ* and Granzyme B secretion.

An antiviral CD4+ T cell response was present in children after ancestral SARS-CoV-2 infection, which was reduced in the youngest age group. We detected significant cross-reactivity of CD4+ T cell responses to the more recently evolved immune-escaping beta variant. Our findings have epidemiologic relevance for children regarding novel viral variants of concern and vaccination efforts.

## Introduction

The SARS-CoV-2 virus appeared in 2019 ^1^ causing a pandemic which, despite effective vaccination, is still a major threat to global health^2^. Children in particular are now facing increasing infection rates due to a reduction of social distancing measures while vaccination of younger age groups has just begun or is not yet available. So far, the disease course in the younger population appears to be mild ^3,4^, however emerging variants may present new challenges. With the current emerging omicron variant hospitalization rates of young children, in particular, appear to be on the rise ^5^. Information on the pediatric immune response after infection or vaccination is of great importance for planning protective strategies in the future. However, data on T cell-mediated immunity in children is still lacking. The identification of antigen-specific T cells via stimulation of patient PBMC with viral peptide pools followed by detection of reactive T cells through activation-induced markers allows the identification and simultaneous phenotyping of these cells using limited available patient material ^6^ and has been broadly used to identify SARS-CoV-2 reactive T cells in adults ^7–14^, and children ^15,16^. The COVID-19 Child Health Investigation of Latent Disease (C19.CHILD) Hamburg Study recruited children from all age groups after the spring 2020 wave of SARS-CoV-2 infections in Hamburg, Germany. Here, PBMC from over fifty SARS-CoV-2 convalescent children, their exposed siblings and unexposed age-matched controls from the C19.CHILD cohort were stimulated with peptide MegaPools (MP) spanning the entire SARS-CoV-2 Spike Glycoprotein of the Wuhan-Hu-1 strain and beta variant as well as predicted peptides representing the remaining entire SARS-CoV-2 Wuhan-Hu-1 strain proteome ^17^ to detect and characterize virus-specific CD4+ T cell responses.

## Methods

### Study cohort and ethics

SARS-CoV-2 convalescent children, exposed seronegative siblings as well as unexposed controls were identified from the COVID-19 Child Health Investigation of Latent Disease (C19.CHILD) Hamburg Study cohort, registered at clinicaltrials.gov (NCT04534608). Briefly, 6113 children (<18 years) who presented voluntarily or were recruited while receiving care in one of the five pediatric hospitals of Hamburg, Germany, were invited for a screening for an acute or recent SARS-CoV-2 infection via PCR and serum antibody testing.

Patients who tested positive in the PCR and/or the antibody screening were invited with all household members for a follow-up appointment, where detailed history was obtained and PCR and serologic SARS-CoV-2 testing were repeated. PBMC samples were obtained from all family members under 18 years. Pediatric unexposed and healthy volunteers, with no known SARS-CoV-2 contact, were welcomed to enrol through the C19.CHILD Study Clinic.

Recruiting was conducted from May 11^th^ until June 30^th^ 2020 after the first infection wave, during and after the first lockdown in Germany. Parents or legal guardians provided written informed consent in all cases. From children over 7 years, consent in writing was obtained whenever possible but also consent in spoken word was accepted. The study was approved by the local ethical committee of Hamburg (reference number: PV7336).

### Serum antibody measurements

For screening purposes, serum samples were tested for SARS-CoV-2 specific antibodies directed against the viral nucleocapsid (IgA/IgM/IgG) using Elecsys® Anti-SARS-CoV-2 Ig assay (Roche) on the cobas e411 system (Roche). Additionally, serum samples were tested for SARS-CoV-2 specific antibodies against the S1 and S2 subunits of the viral Spike protein using LIAISON® SARS-CoV-2 IgG serology test (DiaSorin).

To evaluate serostatus for “common cold” coronaviruses (HCoV) and to further confirm SARS-CoV-2 serostatus, serum antibodies (IgG) against the viral nucleocapsid of HCoV strains 229E, NL63, OC43, HKU1 and SARS-CoV-2 anti - S1 subunit, anti - receptor binding domain as well as anti - nucleocapsid were measured by using the recomLine SARS-CoV-2 IgG® assay (Mikrogen).

Antibody screening was performed IVD conform according to the manufacturer’s instructions. Since patient recruiting was performed in a low prevalence setting, (<5 %), we used an orthogonal testing algorithm. Therefore, samples were considered as SARS-CoV-2 positive if all three tests were positive, negative in three tests was considered as SARS-CoV-2 negative. Participants with discordant results were excluded from later analysis. Unexposed controls were required to report no known SARS-CoV-2 exposure and to be negative in the SARS-CoV-2 PCR test and in the three antibody tests.

### Sample acquisition / processing

Pediatric blood samples (2-10 ml) were collected in EDTA and processed within 24 hours. PBMC were isolated by gradient centrifugation using SepMate tubes® and Lymphoprep® (StemCell) according to the manufacturer’s instructions. PBMC were cryopreserved in freezing medium containing 50% FBS (Capricorn), 30% RPMI (Gibco) and 20% DMSO (AppliChem) and stored in liquid nitrogen until further analysis.

For analysis, frozen aliquots of PBMC were incubated for one minute in a 37°C water bath, and subsequently thawed in prewarmed RPMI by gentle pipetting.

### PBMC peptide stimulation

Peptide stimulation was conducted using previously described ^14,17^ peptide MegaPools optimized for stimulation of CD4+ T cells spanning the entire SARS-CoV-2 spike glycoprotein (Spike-OS_MP) as well as predicted peptides representing the remaining entire SARS-CoV-2 proteome (R_MP) of the first described original strain (Wuhan-Hu-1). Additionally, a peptide pool of overlapping 15-mer by 10 amino acids covering the entire SARS-CoV-2 beta variant spike glycoprotein (Spike-BV_MP) was used.

Thawed PBMC were incubated in 5ml RPMI + human serum (Pan Biotech) 5% + Benzonase (Sigma-Aldrich) 50 U/ml for 1 hour (37°C 5% CO2), followed by a washing step with 15ml RPMI + human serum (HS) 5%. Afterwards all available cells were equally divided to be stimulated for 24 hours (37°C 5% CO2) in 200µl RPMI + HS 5% in 96-well U-bottom plates with mentioned peptide MegaPools (1µg/ml/peptide), PHA-L (Invitrogen) (1µg/ml) as positive control and an equimolar amount of DMSO to serve as negative control. After 24h of stimulation cell culture supernatant was carefully removed and stored at −20°C for later multiplex cytokine analysis. Incubation was stopped by washing cells in PBS. Expression of activation-induced markers (CD69 and OX40) in response to specific peptide stimulation, as well as their memory phenotype were measured by flow cytometry (flow cytometry antibodies are listed in Supplementary Table 1).

### Flow cytometry

After thawing, samples for ex-vivo immune phenotyping were washed twice with PBS, equally split into two aliquots, and analyzed with two optimized panels for investigating T cell, B cell and innate immune cell phenotypes, established previously and slightly adapted ^18^.

The staining procedure for ex-vivo immune phenotyping as well as for PBMC after peptide stimulation was as follows: Cells were stained with Near-IR Dead Cell Stain Kit (Invitrogen) incubated for 15 minutes in the dark at room temperature (RT) and subsequently stained with an antibody cocktail (Supplementary Table 1, Supplementary Figure 1) for 20 minutes at RT. Following washing with PBS, cells were fixed in 1% PFA (Morphisto) for one hour at 4°C, which was removed by PBS wash. Cells were kept at 4°C until acquisition.

Comparability of fluorescence intensities was regularly tested with Rainbow Calibration Particles (BD Sphero). For compensation, Anti-Mouse or Anti-Rat Ig, κ/Negative Control Compensation Particles Set (BD Biosciences) was used for antibodies, and ArC™ Amine Reactive Compensation Beads (Invitrogen) were used for the Dead Cell Stain Kit. Representative gating strategies are shown in Supplementary Figure 1.

Antibody concentrations to achieve optimal separation of targeted populations were evaluated by titration in preliminary experiments using anonymous buffy coats, obtained at the Department of Transfusion Medicine at the University Medical Center Hamburg-Eppendorf from adult blood donors, who provided their written informed consent.

All flow cytometry measurements were performed with a BD FACSymphony A3 flow cytometer in the Cytometry and Cell Sorting Core Unit at University Medical Center Hamburg-Eppendorf.

### Multiplex detection of cytokines

Detection of cytokines in the cell culture supernatant of stimulated cells was performed using LEGENDplex™ Human CD8/NK Panel (13-plex, BioLegend) suitable for detection of IL-2, IL-4, IL-10, IL-6, IL-17A, TNF-α, sFas, sFasL, IFN-γ, Granzyme A, Granzyme B, Perforin, Granulysin, according to the manufacturer’s instructions. Briefly, freshly prepared provided cytokine standard or thawed cell culture supernatant was mixed with cytokine-specific beads, incubated for 2 hours and washed. After sequential incubation of bead-bound cytokines with biotin-labeled cytokine detection antibodies and streptavidin PE antibodies, non-binding antibodies were washed off and PE-labeled bead-bound cytokines were subsequently analyzed by flow cytometry. Quantification of cytokines was carried out using the standard. The data was analyzed using the online LEGENDplex™ Data Analysis Software of the manufacturer. The assay was performed in duplicates and mean values of each sample were used for further analysis.

### Data Analysis and Statistics

Data analysis, graphs and statistics were prepared with FlowJo version 10, and R 4.0.5 (packages: tidyverse, rstatix, splines, emmeans, kableExtra, magrittr, heatmaply). Paired sample analyses were performed by paired t-test. The stimulation index and fold increases showed highly skewed distributions and were therefore log-transformed for further analysis. An unpaired t-test was used for comparing two groups. Categorical variables were compared using Fisher’s exact test. Comparisons between three groups were done with one-way ANOVA and post hoc pairwise t-tests, if the ANOVA-F test was significant, thus following the closed test principle, no adjustment for multiple testing was necessary for pairwise comparisons. The association between age and the stimulation index was explored using a non-parametric spline regression with age and serology group and sex as independent variables. An interaction between spline age and serology group was initially included in the model, but it was removed from the final model, if it did not significantly increase the model fit. A p-value < 0.05 was considered as statistically significant in all analyses. As this study had an exploratory nature, we refrained from adjusting for multiple testing.

For analysis of flow cytometry experiments, samples with less than 5000 acquired alive CD3+ cells (using the DMSO control as reference for peptide stimulations) were excluded from further analysis.

Analysis of AIM+ cells was conducted as described previously ^8,13,14^, by calculating of a stimulation index by dividing the frequency of OX40+CD69+ cells within CD4+ cells after peptide stimulation with the frequency of the same cell subset in the negative control (DMSO). In case no OX40+CD69+ cells were detected with DMSO, the lowest detected frequency for DMSO in the given experimental group was used.

## Results

### Study cohort characteristics

The study cohort consisted of 126 participants: 51 seropositive children as determined by positivity in all of three separate serological tests covering the viral Spike and Nucleocapsid, 24 seronegative siblings living in a shared household with an infected individual and 51 age- and gender-matched unexposed controls. There were no significant differences in the age- and gender distribution of these groups (Table 1). For children whose families were able to give a detailed account of past infection (68 of 75 seropositive children and seronegative siblings), 55% of seropositives and 43% of seronegative siblings reported symptoms consistent with COVID-19. Most families with several symptomatic children could no longer recall the exact date of symptom onset (DSO) for each individual child. However, for 26 out of 35 symptomatic children, families were able to recall DSO of the first symptomatic child in the household which we applied as an approximation to all siblings. The mean time since DSO at the time of sampling was similar within the two groups: 84 days (range 51 – 115 days) for seropositives and 83.4 days (range 62 – 103 days) for seronegative siblings.

**Table 1.** Table showing basic characteristics of the cohort. Number of participants (n) per each analysis and subgroup is indicated as data was available. Groups according to SARS-CoV-2 serology: seropositive, seronegative (exposed) siblings, unexposed controls. The last two columns describe the statistical analyses per row, the p values were calculated with the statistical test as indicated in the last column. HCoV: human “common cold” coronaviruses 229E, NL63, OC43, HKU1. PSO: days post-symptom onset of the first symptomatic child in the family.

|  | Seropositives<br>(n=51) | Seroneg. siblings<br>(n=24) | Unexp. controls<br>(n=51) | p | Test |
| --- | --- | --- | --- | --- | --- |
| Age (years, mean ± range) | 10.63 (0 - 17) | 8.79 (1 - 16) | 10.78 (1 - 17) | 0.170 | ANOVA |
| Sex (female) | 21 (41%) | 12 (50%) | 27 (53%) | 0.523 | Fisher's exact |
| HCoV serology (positive) | 25 (49%) | 8 (33%) | 18 (35%) | 0.291 | Fisher's exact |
|  | Seropositive<br>(n=47) | Seronegative<br>(n=21) |  |  |  |
| Symptoms (yes) | 26 (55%) | 9 (43%) |  | 0.433 | Fisher's exact |
|  | Seropositive<br>(n=21) | Seronegative<br>(n=5) |  |  |  |
| Days PSO (mean ± range) | 84 (51 - 115) | 83.4 (62 - 103) | | 0.907 | t-test |

SARS-CoV-2 seropositive study participants were all convalescent of an ancestral variant, closely related to the original Wuhan-Hu-1 strain, since recruiting took place before the occurrence of the first SARS-CoV-2 variants of concern.

### Antigen-specific CD4+ T cells after infection with ancestral SARS-CoV-2 variant

PBMC from all donors were stimulated with peptide MegaPools derived from the first described original SARS-CoV-2 strain (Wuhan-Hu-1) spanning the entire Spike glycoprotein (Spike-OS_MP) and predicted epitopes from the remaining proteome (R_MP). CD4+ T cell response was measured based on the expression of AIM markers (OX40 and CD69) and compared to a negative control consisting of the peptide mix solvent DMSO at the same concentration as in the peptide mix (Figure 1 A and Supplementary Figure 1 A). When compared to the carrier control (DMSO), stimulation with R_MP led to an increase in the expression of AIM markers in all groups. Increased expression of AIM markers after Spike-OS_MP stimulation was detectable in the seropositive and unexposed control, but not in the seronegative group (Figure 1 B + C). This reflects a combination of antigen-specific CD4+ T cell activation together with an additional unspecific immune activation by peptide stimulation itself over DMSO.

**Figure 1.**
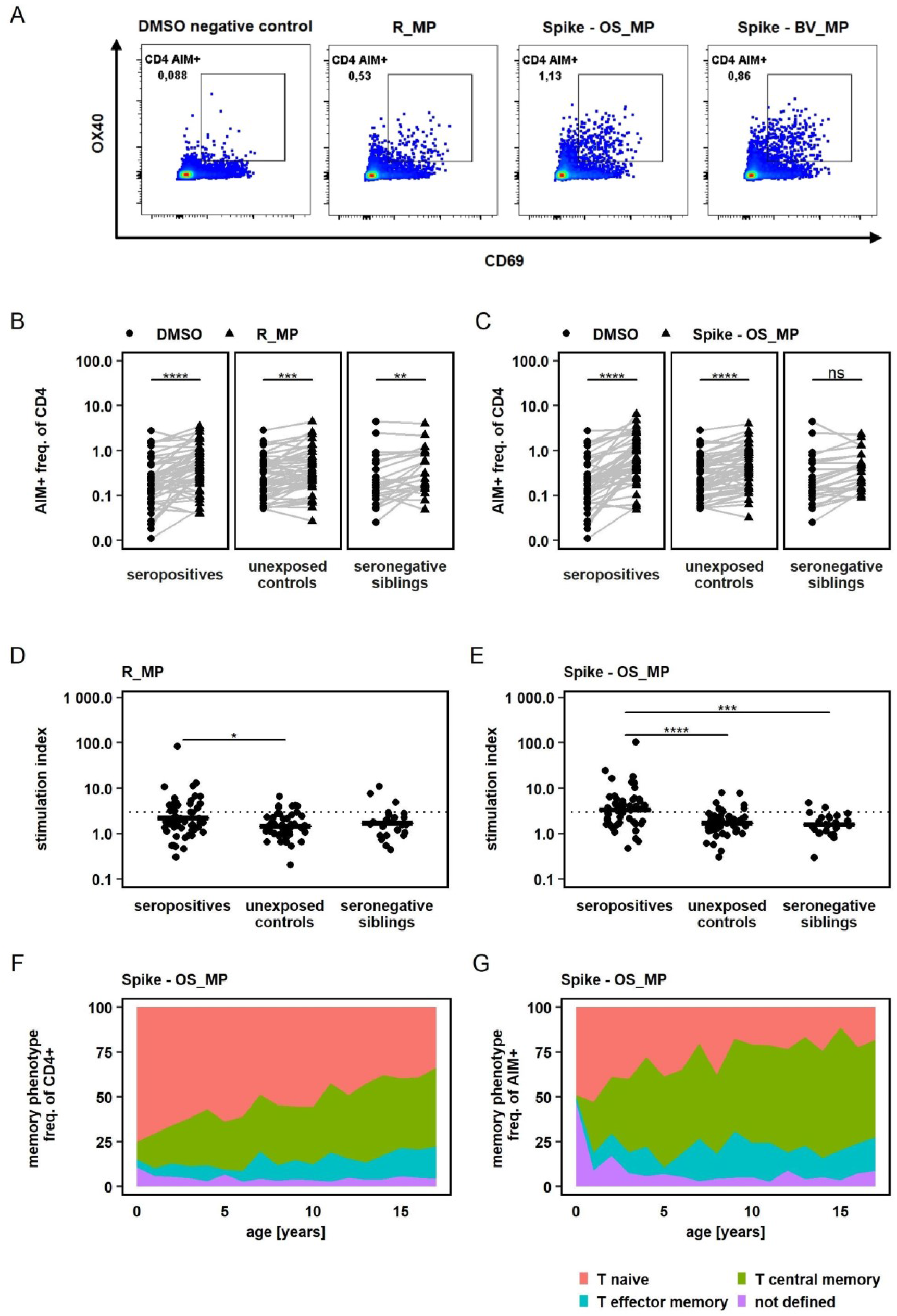
A: Flow cytometry results of a representative SARS-CoV-2 seropositive study participant, gated on alive CD3+ CD4+ lymphocytes. PBMC were equally divided and stimulated over 24h with either DMSO (negative control), PHA-L (positive control) or SARS-CoV-2 derived peptide MegaPools covering the spike glycoprotein of the original strain Wuhan-Hu-1 (Spike-OS_MP), beta variant (Spike-BV_MP) or the remaining original strain’s proteome (R_MP). CD4 AIM+ T cells were identified by their CD69 and OX40 expression. CD4 AIM+ gate was set equal for all samples of all participants after comparability of fluorescence intensities was assured by rainbow bead calibration. B + C: Frequency of AIM+ cells, determined by CD69 and OX40 expression, after stimulation with R_MP and Spike - OS_MP in comparison to AIM+ frequency after DMSO exposure. Study groups were determined by their SARS-CoV-2 serostatus and SARS-CoV-2 exposure. Paired t – tests were used to determine P values. D+ E: Comparison of T cell response towards peptide stimulation between study groups. T cell response was quantified by using a Stimulation Index (SI) which was calculated by dividing the freq. of AIM+ cells after peptide stimulation by the freq. of AIM+ cells of the DMSO negative control. A SI > 3 (dashed line) was defined as response. Statistical comparisons were carried out by one way ANOVA and post hoc pairwise t – tests. F + G: Memory phenotype of total CD4+ and AIM+CD4+ T cells after peptide stimulation with Spike - OS_MP. Mean values of all study participants irrespective of SARS-CoV-2 serostatus are displayed *P < 0.05, **P < 0.01, ***P < 0.001, ****P < 0.0001, ns - not significant

To account for this unspecific immune activation and to be able to compare between groups, we used the stimulation index (SI), calculated as previously reported ^8,13,14^ as individual MP response divided by DMSO response. SI in the seropositive group was increased over unexposed controls for R_MP as well as Spike-OS_MP stimulation. Additionally, on Spike-OS_MP stimulation SI was increased in seropositives over seronegative siblings (Figure 1 D + E). By applying a SI threshold of > 3 to define responders ^8,13,14^, 61 % (31 of 51) of seropositive children showed a specific CD4+ T cell response to either Spike-OS_MP or R_MP stimulation, with 55 % (28 of 51) being responsive to Spike-OS_MP and 33 % (17 of 51) to R_MP. This provides further evidence for the generation of antigen-specific T cells after SARS-CoV-2 infection in children and additionally underscores the dominant immune response elicited by the SARS-CoV-2 Spike protein.

Additionally, reactive T cells were detected in 8 % (2 of 24 Spike-OS_MP) and 13 % (3 of 23 R-MP) of seronegative siblings, as well as in 14 % (7 of 51 Spike-OS_MP) and 12 % (6 of 51 R_MP) of unexposed controls.

Without detectable differences in T cell response between seronegative siblings and unexposed controls, our data indicate that the intensity of exposure to SARS-CoV-2 in household members, which did not lead to a humoral immune response (seronegative siblings) was generally also not sufficient to induce a systemic CD4+ T cell response.

Naive versus memory phenotype of AIM+ CD4+ T cells was determined using CD27 and CD45RA expression (Supplementary Figure 1 A). This revealed that SARS-CoV-2 specific AIM+ CD4+ T cells predominantly exhibited a central memory phenotype. The proportion of naive cells within the AIM+ fraction was, however, higher in younger children paralleling the higher naive fraction in total CD4+ cells (Spike – OS_MP in Figure 1 F + G, R_MP and Spike – BV_MP in Supplementary Figure 2).

### Cross-reactivity to SARS-CoV-2 beta variant after infection with the ancestral variant

Occurrence of new SARS-CoV-2 variants of concern, characterized by higher transmissibility and a certain immune escape has changed the pandemic’s dynamic several times, reviewed in ^19^. It is therefore of great interest whether T cell responses to earlier SARS-CoV-2 infections or the current vaccinations approved in children, which are based on the original Wuhan-Hu-1 strain by either containing inactivated virus^20,21^ or the Spike protein sequence^22,23^, lead to generation of cross-reactive CD4+ T cells against new SARS-CoV-2 variants.

We evaluated the T cell response towards the beta variant, a WHO variant of concern due to its capability to escape humoral immunity ^24^, in archived pediatric samples after infection with an ancestral SARS-CoV-2 variant. PBMC were stimulated with a peptide MegaPool of overlapping peptides spanning the entire Spike glycoprotein of the beta variant (Spike-BV_MP). This resulted in a significant increase of CD4+ AIM markers in the seropositive cohort (Figure 2 A). 45 % of seropositives (23 of 51), 12 % of unexposed controls (6 of 51) and 21 % of seronegative siblings (5 of 24) could be identified as responders (SI >3). SI was significantly higher in seropositive individuals when compared to unexposed controls and seronegative siblings (Figure 2 B). Our results suggest that prior infection with an ancestral SARS-CoV-2 variant is associated with the generation of T cells showing reactivity against the immune escaping SARS-CoV-2 beta variant in a relevant proportion of children.

**Figure 2.**
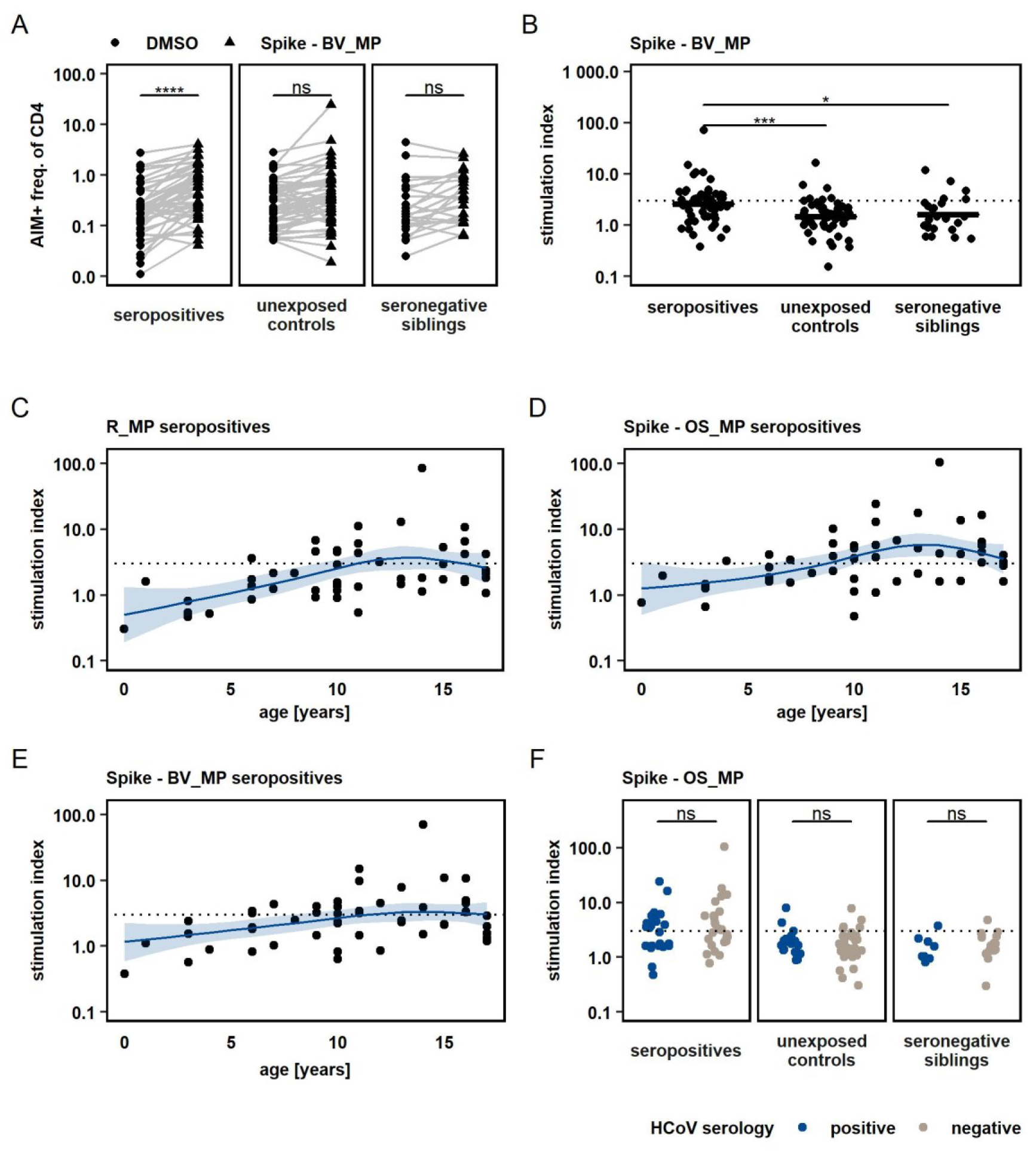
A: Frequency of AIM+ cells after stimulating PBMC with Spike – BV_MP and DMSO negative control. Paired t – tests were used to quantify P – values. B: Comparison of T cell response, quantified by stimulation index, to Spike – BV_MP peptide pool between study groups. Statistical analysis was performed with one way ANOVA followed by pairwise t – tests. SI > 3 (dashed line) was defined as response. C + D + E T cell responses upon peptide stimulation as quantified by stimulation index are displayed according to the participants’ age. Each dot represents the T cell response of an individual seropositive study participant. The respective peptide pool is indicated on top of each graph. The effect of age on the magnitude of T cell response, was further analyzed with a non-parametric multivariate regression analysis, by using a spline model (blue lines with light blue areas indicating 95% confidence intervals). F: T cell responses towards Spike – OS_MP stimulation were compared between participants with different serostatus for “common cold” coronaviruses (HCoV, strains 229E, NL63, OC43 and HKU1). Analyses were conducted within study groups, which were defined by SARS-CoV-2 exposure and serostatus. Unpaired t – test was used to quantify P values. *P < 0.05, **P < 0.01, ***P < 0.001, ****P < 0.0001, ns - not significant

### Lower antigen-specific T cell detection rate in very young children

We analyzed the effect of age on SARS-CoV-2 specific CD4+ T cell responses (Figure 2 C,D,E), by applying a non-parametric spline model to fit the data on T cell responses as measured by SI plotted by age in years. This model predicts an increase in antigen specific CD4+ T cell response with age, until a plateau is reached between ten and fifteen years. Also, we detected an impaired capacity to mount specific T cell responses (SI > 3) in preschool children using all tested SARS-CoV-2-derived peptide pools. At the same time, we did not detect an overall reduced capacity of CD4+ T cells in younger children to express AIMs upon activation in the positive control (PHA, Supplementary Figure 4).

### Influence of prior HCoV infections

To evaluate the possible influence of T cell cross reactivity elicited by prior infections with common cold Coronavirus (HCoV), we assessed the serostatus of HCoV strains 229E, NL63, OC43 and HKU1 in all study participants. 40 % of the participants in the study were identified as seropositive for at least one of the tested strains. When comparing T cell response quantified as SI for stimulation with R_MP, Spike – OS_MP and Spike – BV_MP between participants with positive and negative HCoV serology, no difference could be seen regardless of SARS-CoV-2 serostatus (Spike – OS_MP Figure 2 F blue and grey, R_MP and Spike – BV_MP in Supplementary Figure 3). Some, but not all unexposed controls and seronegative siblings with SI > 3 in any of the peptide pools had a positive HCoV serology. This indicates that the identified SARS-CoV-2 reactive T cells in seronegative siblings and unexposed controls cannot be explained only by cross reactive T cell memory from prior HCoV infections.

### Cytokine profile of SARS-CoV-2-reactive T cells

To further characterize the T cell response, thirteen different cytokines were measured in culture supernatants after exposure of PBMC to SARS-CoV-2 peptide MegaPools. We could detect increased cytokine levels for nearly all of the quantified cytokines after peptide stimulation when compared to DMSO negative control (data not shown). To control for unspecific or bystander immune activation and to be able to attribute cytokine secretion to a specific T cell response, we calculated fold increases in cytokine concentration after peptide stimulation over cytokine concentration in corresponding DMSO negative controls analogous to the stimulation index for T cell response. These were compared between seropositive children with a clear T cell response (SI > 3 as quantified using activation induced markers) and unexposed controls and seropositive children both without a clear T cell response (SI < 3).

The analysis revealed levels of IFN-γ and Granzyme B, both associated with viral defense, were consistently higher in seropositive T cell responders after Spike-OS_MP, R_MP and Spike-BV_MP stimulation compared seropositives and unexposed controls both without a T cell response (Figure 3 A – F). Since PBMC were stimulated in bulk, the elevated IFN-γ and especially Granzyme B levels could indicate concomitant CD8+ activation.

**Figure 3.**
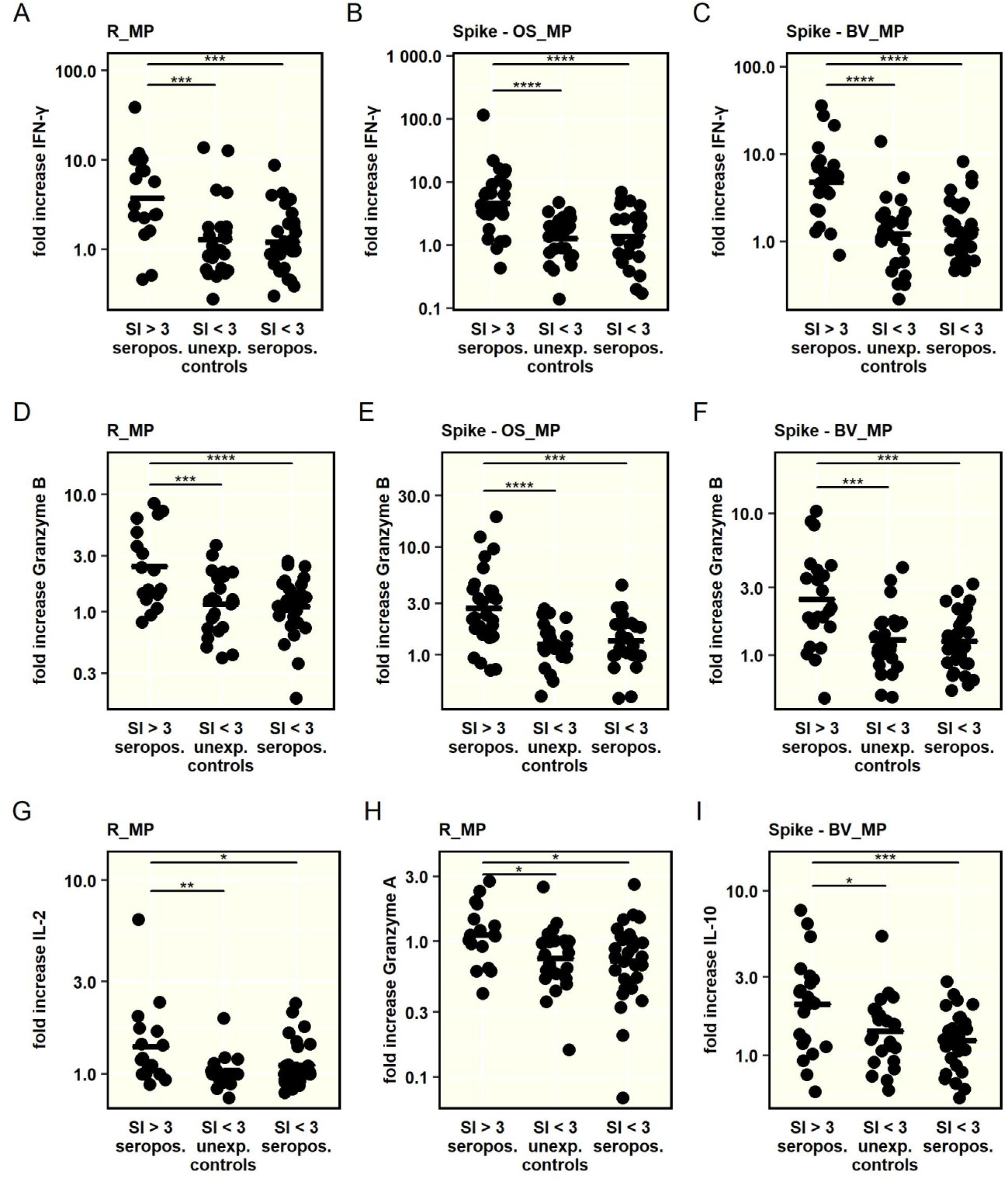
The concentration of 13 cytokines was determined in cell culture supernatants after stimulation with SARS-CoV-2-derived peptide MegaPools. The respective peptide pool, used for stimulation, is indicated on top of each graph. Comparison was conducted between seropositive children with and without a clear T cell response and unexposed controls without a T cell response. T cell response was defined as a stimulation index (SI) > 3 in the AIM assay upon peptide stimulation A – I: Comparison of IFN-γ, Granzyme B, IL-2, Granzyme A and IL-10 secretion between groups. To adjust for unspecific cytokine secretion, comparison was performed by using the fold increase in cytokine concentration after peptide stimulation over cytokine concentration in DMSO treated samples. One way ANOVA and post hoc pairwise t – tests were used to quantify P values. *P < 0.05, **P < 0.01, ***P < 0.001, ****P < 0.0001, not significant – not displayed

In seropositive T cell responders, R_MP stimulation led to an increased secretion of IL-2 (Figure 3 G) and Granzyme A (Figure 3 H). Interestingly, the beta-variant-based Spike-BV_MP was the only peptide pool eliciting the secretion of anti-inflammatory IL-10 in association with a CD4+ T cell response (Figure 3 I, IL-2 and Granzyme A response not shown for Spike-OS_MP and Spike-BV_MP. IL-10 not shown for Spike-OS_MP and R_MP).

### No long-term alterations of the immune phenotype in convalescent pediatric patients

Respiratory infections are known to elicit long-term changes in the phenotype of innate and adaptive immune cell populations ^25,26^. Therefore, we performed a broad immunologic phenotyping of study participants in parallel PBMC samples using two flow cytomery panels for a detailed T cell profiling and quantification of main immune cell subsets, respectively (Supplementary Figure 1 B and C). We found that the relative abundance of T-, B- and innate cell subsets was similar in seropositive children, seronegative siblings and unexposed controls (Figure 4 A and B). Thus, in this pediatric cohort, no long-term immune phenotypic changes after SARS-CoV-2 infection or exposure could be demonstrated.

**Figure 4.**
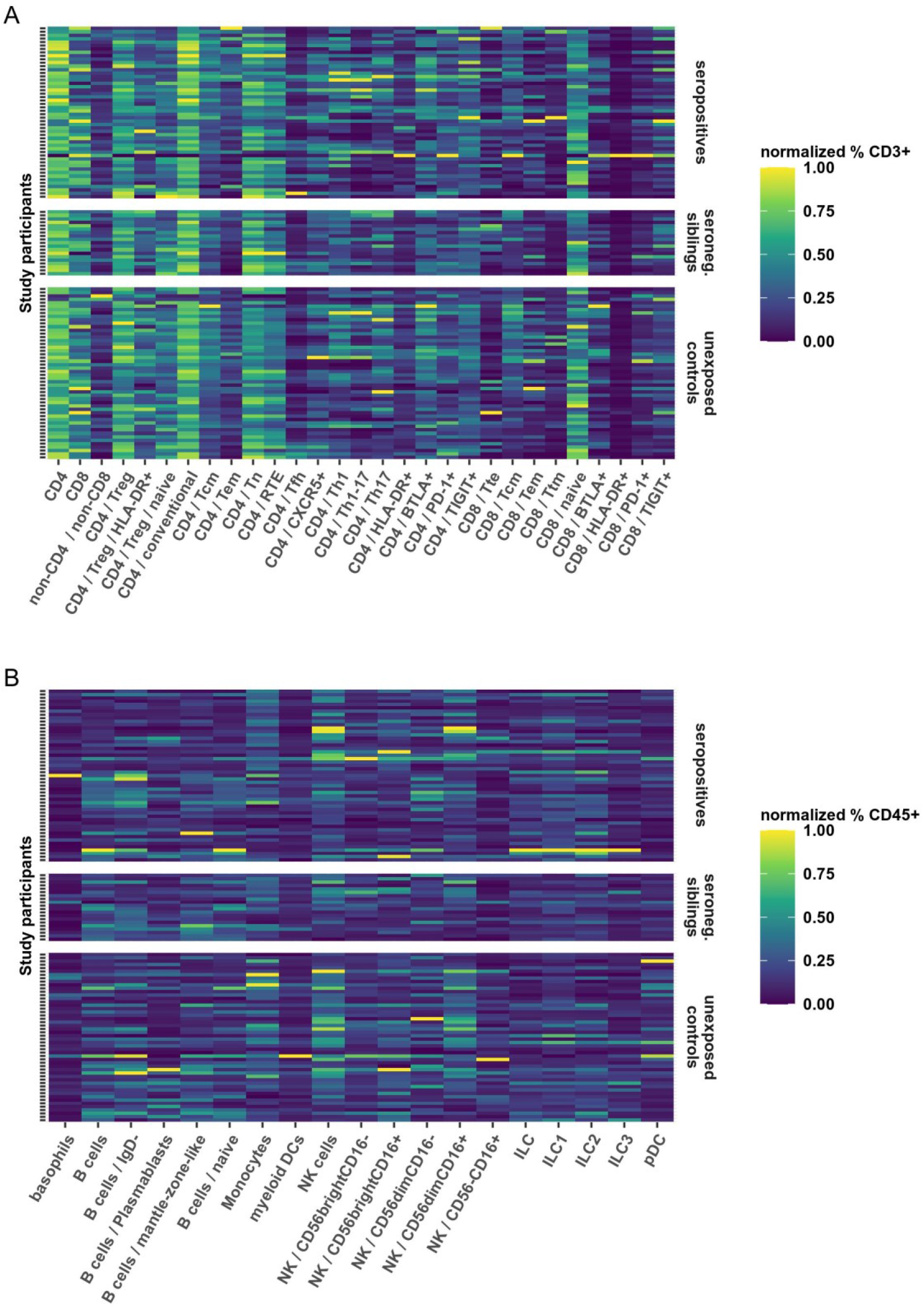
**Heatmaps** showing the relative frequency of **T cell subsets** (**A**) as frequency of CD3 positive live cells and **B- and innate cell subsets** (**B**) as frequency of CD45 positive live cells. Further subdivision of subsets is indicated by forward slash “/”, whereby the parent population is indicated in front of the child population. The frequencies have been normalized for each cellular subset to utilize the full color scale, which is indicated to the right of both panels. Each row represents a PBMC sample from a single participant. Samples are grouped according to seroconversion status (right y axis). Furthermore, samples within groups are sorted vertically according to the age of the participant, beginning with the oldest (top) to the youngest (bottom). MAIT: Mucosa Associated Invariant T cells, Treg: regulatory T cells, Tcm: central memory T cells, Tem: effector memory T cells, Tn: naive T cells, RTE: recent thymic emigrants, Tfh: T follicular helper cells, Th: T helper cells, HLA: human leukocyte antigen, BTLA: B- and T-lymphocyte attenuator, PD-1: programmed death 1, TIGIT: T cell immunoreceptor with Ig and ITIM domains, Tte: terminally differentiated effector T cell, Ttm: transitional memory T cell, pDC: plasmacytoid dendritic cells, NK: natural killer cells, ILC: innate lymphoid cells.

## Discussion

Antigen specific CD4+ T cells play a central role in the immune response to a viral infection, as they may reduce disease severity after re-infection by a more rapid clearance of the virus and better overall disease control, as reviewed in ^27^. The presence of antigen specific T cells after SARS-CoV-2 infection in adult cohorts has been widely demonstrated ^7–10,14,28–30^. Here, we analyzed the T cell response in a large cohort of SARS-CoV-2 convalescent children, their seronegative siblings as well as unexposed controls. We provided further evidence that a high proportion of children who seroconverted to SARS-CoV-2 were able to mount specific CD4+ T cell responses, that were still detectable up to over 100 days after infection. In contrast to adult cohorts ^31^ we could identify specific T cell responses in only a minority of SARS-CoV-2 exposed but not seroconverted children (seronegative siblings). In our pediatric cohort we showed a T cell response attributable to a prior SARS-CoV-2 infection in 61% of seropositive children, which is less than the reported response rates of 77 – 100% of acutely ill ^7,10,14^ and 81 – 100% of convalescent adult patients ^8,10,12,29,30^. These findings are in line with previous studies investigating systemic T cell responses in children after respiratory infections like SARS-CoV-2 ^15,16^ or Influenza ^32^.

Until now data on pediatric T cell response after SARS-CoV-2 infection is scarce with cohort sizes not allowing for analysis over different age groups ^15,16^. Importantly, we showed an increase in SARS-CoV-2-specific CD4+ T cell responses by age within this pediatric cohort. Reduced capacity to mount specific T cell responses was particularly seen in preschool children. A comparable age-effect showing lower T cell responses after SARS-CoV-2-infection in children than adults has previously been demonstrated ^16^. Such age related differences have been reported, especially in infants, regarding the interplay between innate and adaptive immunity and the development of memory T cell responses including a lower inflammatory reaction upon pathogen challenge and vaccination ^33,34^.

We provided functional data on the phenotype of the pediatric CD4+ T cell response to SARS-CoV-2 in children showing a strong antiviral response and provide indirect evidence for a cytotoxic CD8+ response involving Granzyme B production. Predominant CD4+ T cell response in convalescent children is likely of Th1 bias (IFN-*γ*).

While a large proportion of seroconverted children appeared to mount strong T cell responses, long-term changes in the immunological phenotype of innate and adaptive cell populations seem not to affect children after SARS-CoV-2 infection. Notably, children in this cohort were typically sampled about three months after COVID-19, with only a minority having exhibited symptoms of infection in the first place. These characteristics may explain the quick normalization of any immunological changes post infection in this cohort. Conversely, in cohorts with a larger proportion of symptomatic or seriously ill children, also CD4+ T cell reactivity could be higher.

Cross-reactive T cells have the potential to reduce disease severity of infections with new viral strains, reviewed in ^35,36^. The presence of SARS-CoV-2 reactive CD4+ T cells in unexposed individuals could be shown ^7,12,14,29,30^ and cross reactivity of CD4+ T cells to epitopes of HCoVs and SARS-CoV-2 was demonstrated ^37^. We analyzed the influence of prior HCoV infections on pediatric SARS-CoV-2 T cell response. In our cohort, we could not detect a difference in the CD4+ T cell response (quantified by SI) upon SARS-CoV-2-derived peptide stimulation between HCoV seropositive and seronegative individuals. Also, SARS-CoV-2-reactive T cells in unexposed controls were only present in a fraction of cases associated with a positive HCoV serology. We observed a seropositivity for HCoV in 49% of SARS-CoV-2 seropositive and 33% of SARS-CoV-2 exposed participants while no severe disease courses were reported in our cohort. It would follow, that the comparatively mild course of COVID-19 in children is not mainly or exclusively explained by their more frequent or recent exposures to related human coronaviruses. More likely, reduced disease severity and lower T cell responses in children are offset by an enhanced capability of innate, tissue resident immune responses in the upper airways resulting in early SARS-CoV-2 control in this age group ^38^.

Importantly, by making use of our relatively large cohort of archived pediatric samples, we demonstrated for the first time to our knowledge a cross reactivity of T cells after infection with an ancestral SARS-CoV-2 variant to an immune escaping SARS-CoV-2 variant of concern (B.1.351-beta variant) in a pediatric population. This adds on to previous findings, describing cross reactive T cells to B.1.351, B.1.1.7 (alpha-variant), B.1.617.2 (delta-variant) and B.1.1.529 (omicron-variant) in adult cohorts after SARS-CoV-2 infection or vaccination ^30,39–41^. T cell response to the beta variant may serve as a raw model for immune escaping variants, as immune escape is mainly attributed to its mutation on position E484 within the spike domain, which is associated with reduced affinity of neutralizing antibodies ^42^ and mutations on this site are also found in SARS-CoV-2 strains B.1.617.2 (gamma), B.1.621 (mu) or B.1.1.529 (omicron) ^43^.

Our data provide important evidence for a certain cross reactivity of the pediatric CD4+ T cell response after infection with an ancestral SARS-CoV-2 variant to mutations in the Spike domain. Current SARS-CoV-2 vaccines approved for use in children, consisting of inactivated virus^20,21^ or the genetic information of the Spike protein ^22,23^, are based on the original SARS-CoV-2 Wuhan-Hu-1 strain. Based on our data generated by stimulation of archived samples from the first wave with peptide MegaPools derived from the later evolved beta variant, we can hypothesize that these vaccines should still elicit adequate T cell responses to emerging immune escaping variants in children. CD4+ T cell responses in preschool children were reduced, which may be offset by antibody responses ^44^ or increased innate responses ^38^. Nonetheless, the role of a lower CD4+ memory T cell response in the youngest should be considered when planning vaccination strategies and if emerging viral variants cause a rise in pediatric disease burden. Notably, first data from the United States indicate an increased hospitalization rate especially in this age group facing the current omicron variant ^5^.

Our study had several limitations. As children were recruited weeks or months after infection, no PCR based confirmation from the acute phase of infection was available. As previous infection was defined through seropositivity alone, a combined positivity in three independent serological tests was required and this approach should minimize the possibility of false positives. A further limitation based on the retrospective study design is that a detailed history from the time of infection as well as the exact time of infection could only be collected for a fraction of participants. Because of a reluctance in small children and their caregivers, as well as the technical difficulty regarding blood draws, younger age groups were relatively underrepresented. Also, CD8+ T cell responses were not investigated - these could provide important insights into cellular anti-SARS-CoV-2 immunity and should be the focus of further studies.

## Conclusion

Here we showed that pediatric CD4+ T cell responses after infection with an ancestral SARS-CoV-2 variant are age dependent, with reduced capability of the youngest to mount specific responses. Antigen specific T cells persist over three months after infection and are cross reactive with the SARS-CoV-2 variant of concern B.1.351-beta variant. We detected a strong antiviral cytokine response in association with SARS-CoV-2-specific T cell activation. Our findings have relevance when planning rational vaccination of children as well as social distancing measures involving the pediatric population in case of emerging SARS-CoV-2 variants.

## Data Availability

All data produced in the present study are available upon reasonable request to the authors

## Acknowledgements

The C19.CHILD Hamburg Study received funding from the Senate Chancellery of the Free and Hanseatic City of Hamburg. The following foundations and organizations have provided financial support: Carlsen Verlag, Dr. Melitta Berkemann Stiftung, Fördergemeinschaft Kinderkrebs-Zentrum Hamburg e.V., Freunde der Kinderklinik des UK Eppendorf e.V., HSV Fussball AG, Joachim-Herz-Stiftung, Michael Otto Stiftung, Michael Stich Stiftung, Nutricia, Stiftung KinderHerz, EAGLES Charity Golf Club e.V., DAMP Stiftung, Kroschke Stiftung, ZEIT-Stiftung.

## Author Contributions

KP, SWG and GAD conceived the study. KP, FS and GAD performed T cell stimulation assays, cytokine analysis and immune phenotyping. ML supervised serology testing. KP, FS, DW, MB, AS, ET, JSzW, ML, SWG and GAD contributed to the experiment design and methodology. JO, TSM, ACM, SWG, GAD were responsible for study supervision and funding. KP, MW, KH, AK, DEZ, LH, MKD, JO, TSM, ACM, GAD coordinated the study cohort and acquired patient samples. Sample preparation was supervised by KP, FS and GAD. KP, GAD, LG, ET, EV, and AZ performed data- and statistical analysis and prepared graphs. All authors provided conceptual input, KP, FS and GAD wrote the manuscript, which was critically revised by all authors.

## Supplementary Material

**Supplementary Table 1.**
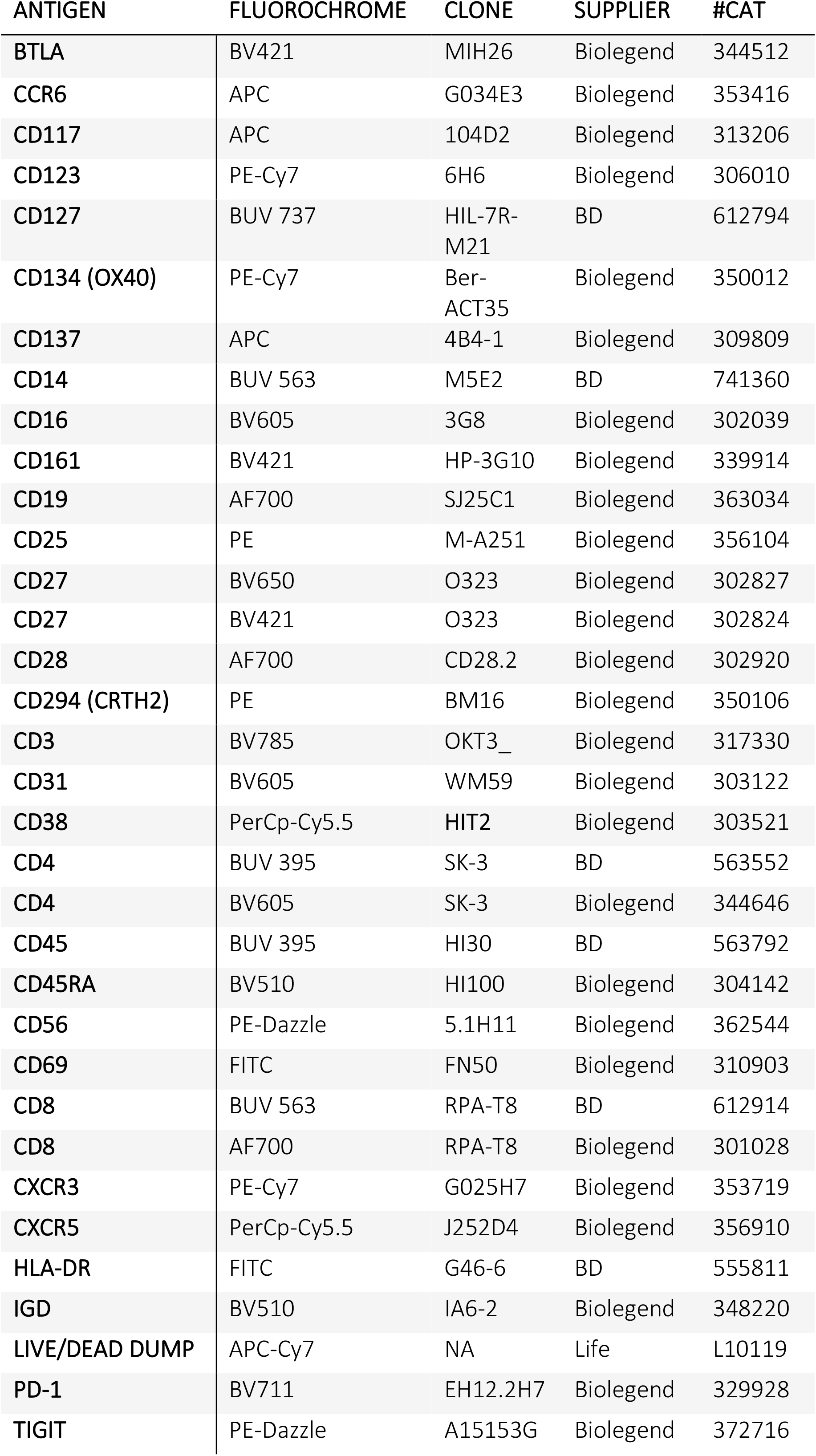
Flow cytometry reagent list.

| ANTIGEN | FLUOROCHROME | CLONE | SUPPLIER | #CAT |
| --- | --- | --- | --- | --- |
| BTLA | BV421 | MIH26 | Biolegend | 344512 |
| CCR6 | APC | G034E3 | Biolegend | 353416 |
| CD117 | APC | 104D2 | Biolegend | 313206 |
| CD123 | PE-Cy7 | 6H6 | Biolegend | 306010 |
| CD127 | BUV 737 | HIL-7R-M21 | BD | 612794 |
| CD134 (OX40) | PE-Cy7 | Ber-ACT35 | Biolegend | 350012 |
| CD137 | APC | 4B4-1 | Biolegend | 309809 |
| CD14 | BUV 563 | M5E2 | BD | 741360 |
| CD16 | BV605 | 3G8 | Biolegend | 302039 |
| CD161 | BV421 | HP-3G10 | Biolegend | 339914 |
| CD19 | AF700 | SJ25C1 | Biolegend | 363034 |
| CD25 | PE | M-A251 | Biolegend | 356104 |
| CD27 | BV650 | O323 | Biolegend | 302827 |
| CD27 | BV421 | O323 | Biolegend | 302824 |
| CD28 | AF700 | CD28.2 | Biolegend | 302920 |
| CD294 (CRTH2) | PE | BM16 | Biolegend | 350106 |
| CD3 | BV785 | OKT3_ | Biolegend | 317330 |
| CD31 | BV605 | WM59 | Biolegend | 303122 |
| CD38 | PerCp-Cy5.5 | HIT2 | Biolegend | 303521 |
| CD4 | BUV 395 | SK-3 | BD | 563552 |
| CD4 | BV605 | SK-3 | Biolegend | 344646 |
| CD45 | BUV 395 | HI30 | BD | 563792 |
| CD45RA | BV510 | HI100 | Biolegend | 304142 |
| CD56 | PE-Dazzle | 5.1H11 | Biolegend | 362544 |
| CD69 | FITC | FN50 | Biolegend | 310903 |
| CD8 | BUV 563 | RPA-T8 | BD | 612914 |
| CD8 | AF700 | RPA-T8 | Biolegend | 301028 |
| CXCR3 | PE-Cy7 | G025H7 | Biolegend | 353719 |
| CXCR5 | PerCp-Cy5.5 | J252D4 | Biolegend | 356910 |
| HLA-DR | FITC | G46-6 | BD | 555811 |
| IGD | BV510 | IA6-2 | Biolegend | 348220 |
| LIVE/DEAD DUMP | APC-Cy7 | NA | Life | L10119 |
| PD-1 | BV711 | EH12.2H7 | Biolegend | 329928 |
| TIGIT | PE-Dazzle | A15153G | Biolegend | 372716 |

**Supplementary Figure 1A.**
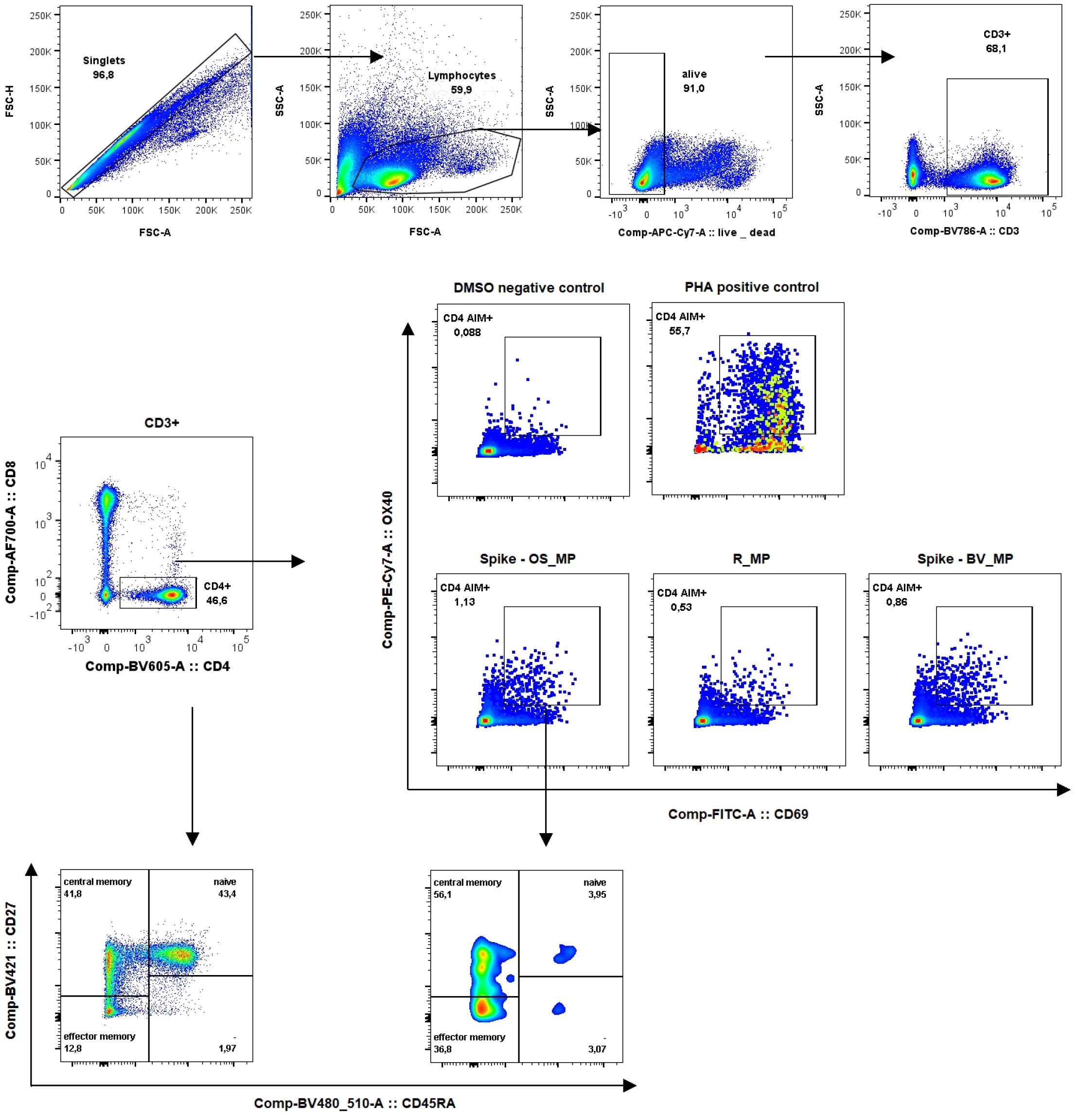
Gating strategy for activation induced markers “Panel AIM” Manual gating strategy for the gating of activation induced markers. Names of subpopulations are shown along with frequency within the parent population. The parent population is indicated above the plots or by arrows showing backgating. For stimulation, the different conditions (negative, positive controls or peptide megapools) are indicated above the plots.

**Supplementary Figure 1B.**
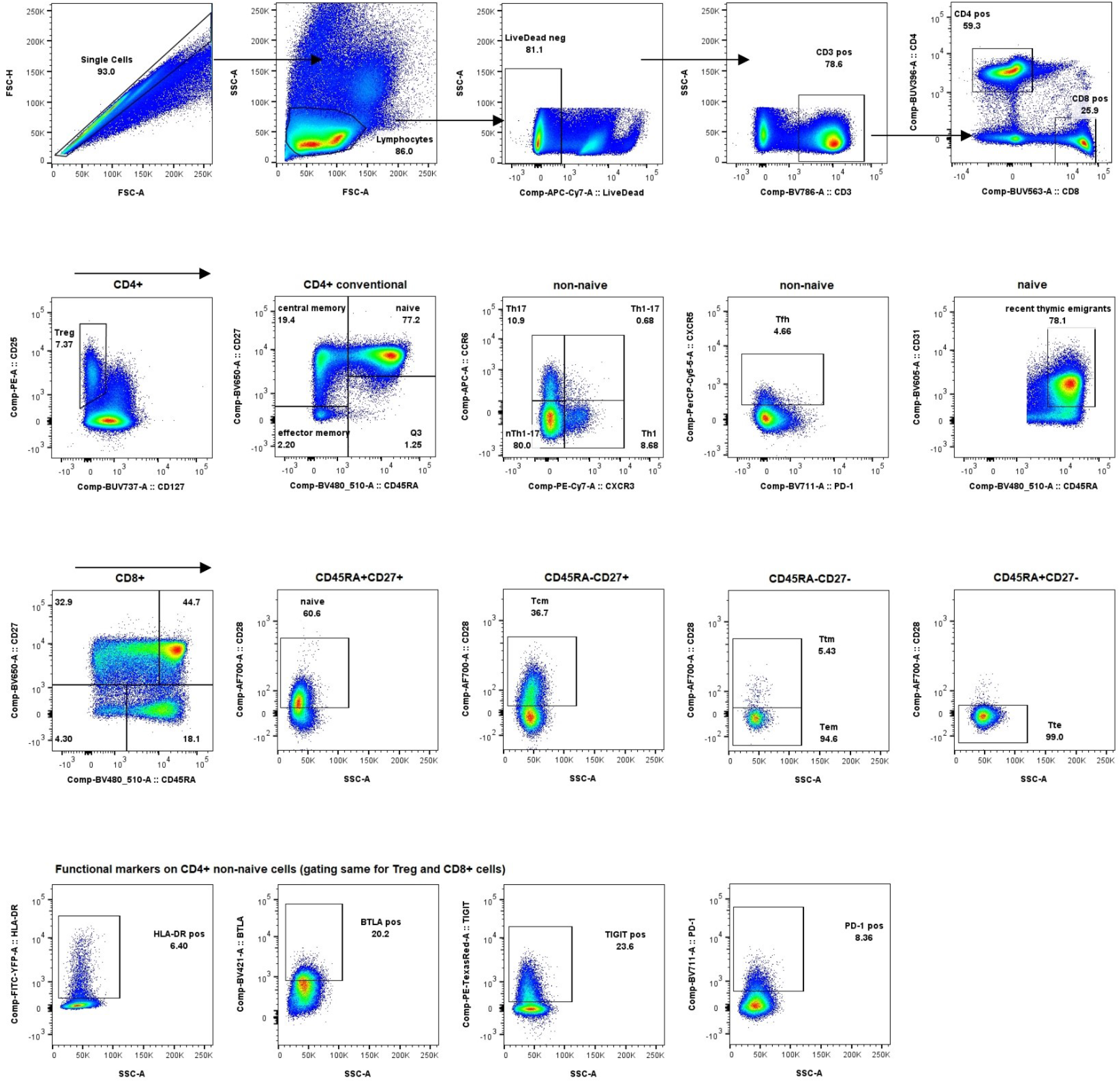
Gating strategy for immune phenotyping, “T cells” Manual gating strategy for T cell subpopulations on a representative sample. Names of subpopulations are shown along with frequency within the parent population. The parent population is indicated above the plots or by arrows showing backgating.

**Supplementary Figure 1C.**
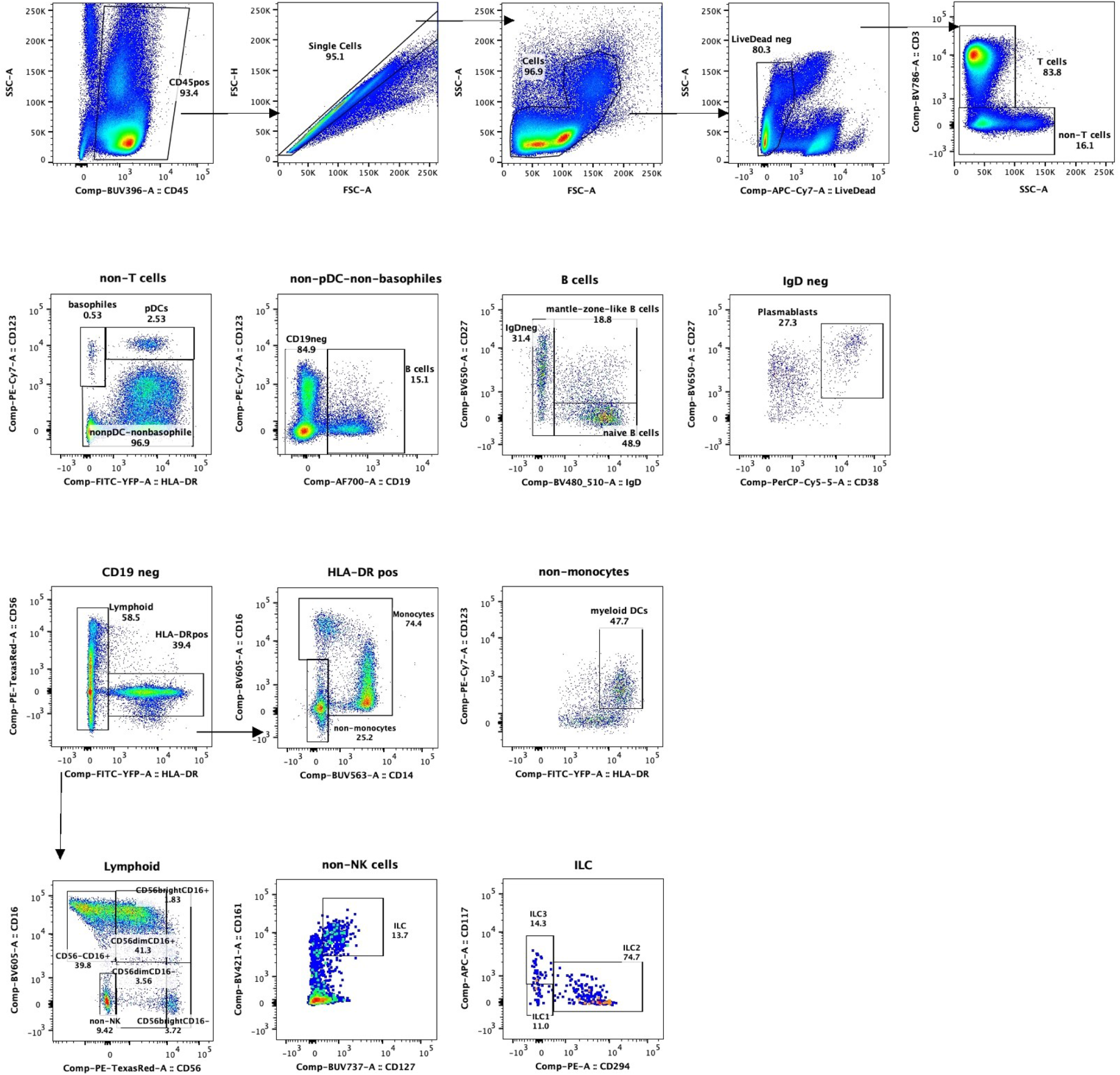
Gating strategy for immune phenotyping, “B cells and innate immune cells” Manual gating strategy for B cells and innate immune cells on a representative sample. Names of sub-populations are shown along with frequency within the parent population. The parent population is indicated above the plots or by arrows showing backgating.

**Supplementary Figure 2.**
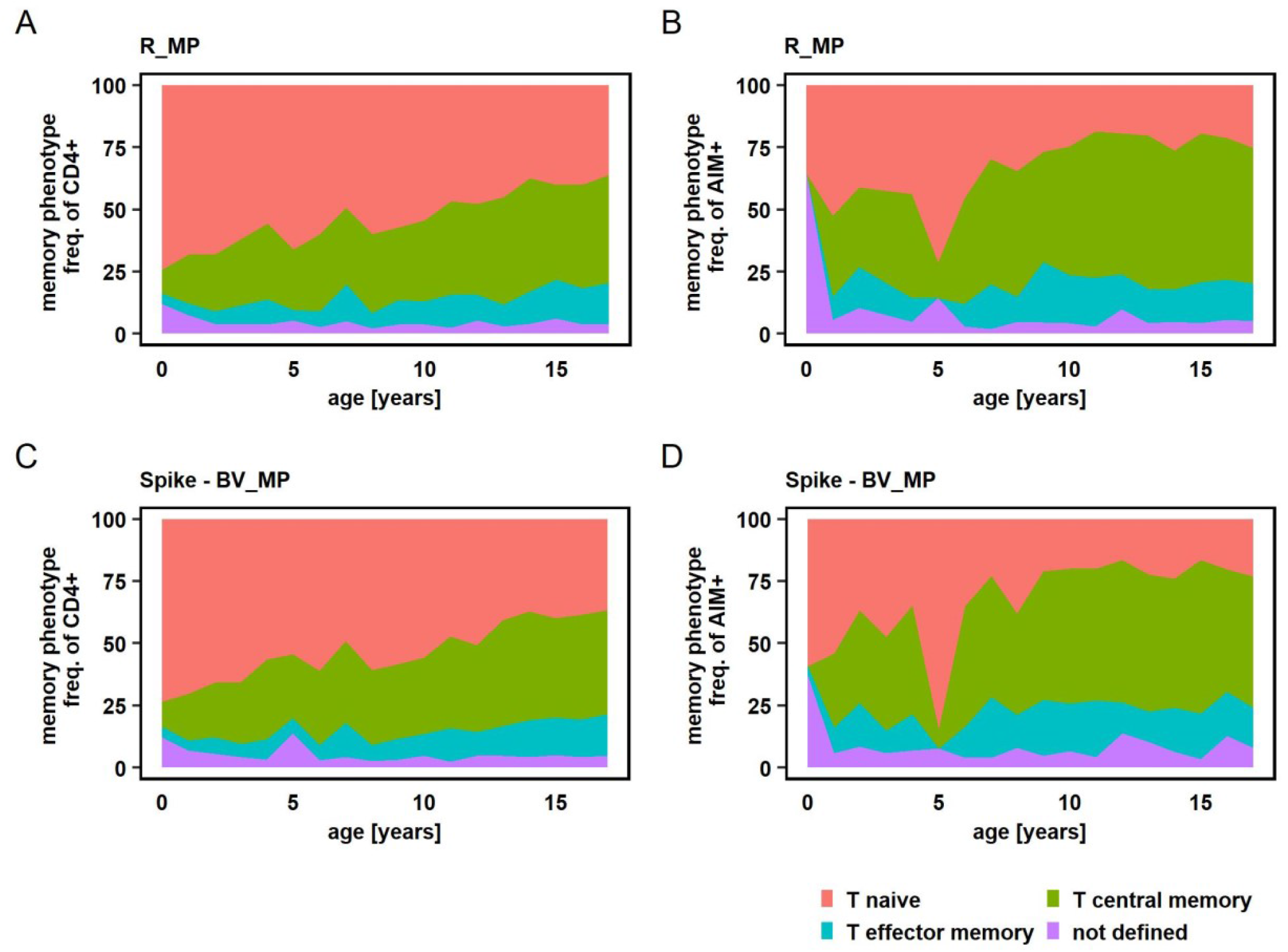
Memory phenotypes upon peptide stimulation Memory phenotypes of total CD4+ and AIM+CD4+ T cells after peptide stimulation with R_MP (A + B) and Spike - BV_MP (C + D). Mean values of all study participants irrespective of SARS-CoV-2 serostatus are displayed.

**Supplementary Figure 3.**
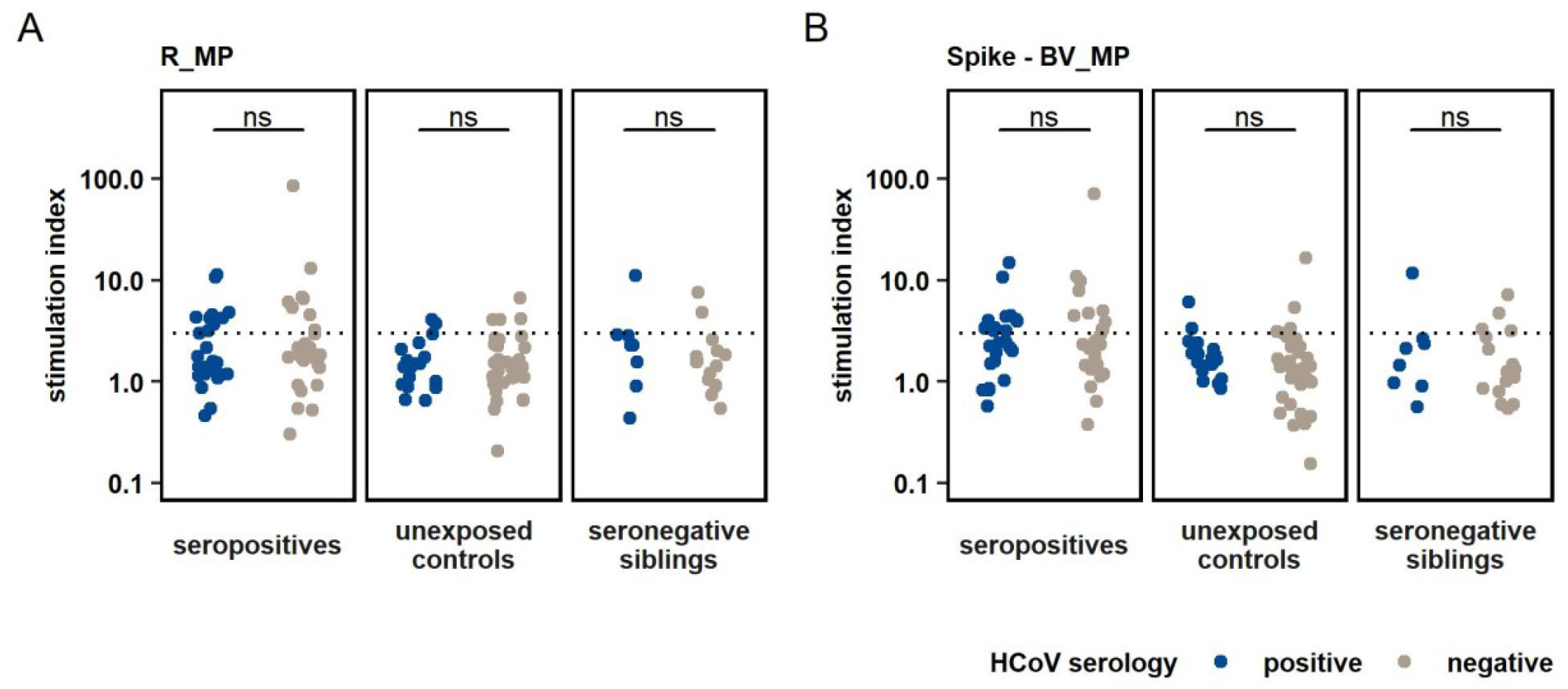
T cell response to peptide stimulation compared between different HCoV serostatus within study groups Comparison of T cell response towards R_MP and Spike – BV_MP stimulation between their different serostatus for “common cold” coronaviruses (HCoV). Analyses were conducted within study groups, which were defined by SARS-CoV-2 exposure and serostatus. Unpaired t – test was used to quantify P values. ns - not significant.

**Supplementary Figure 4.**
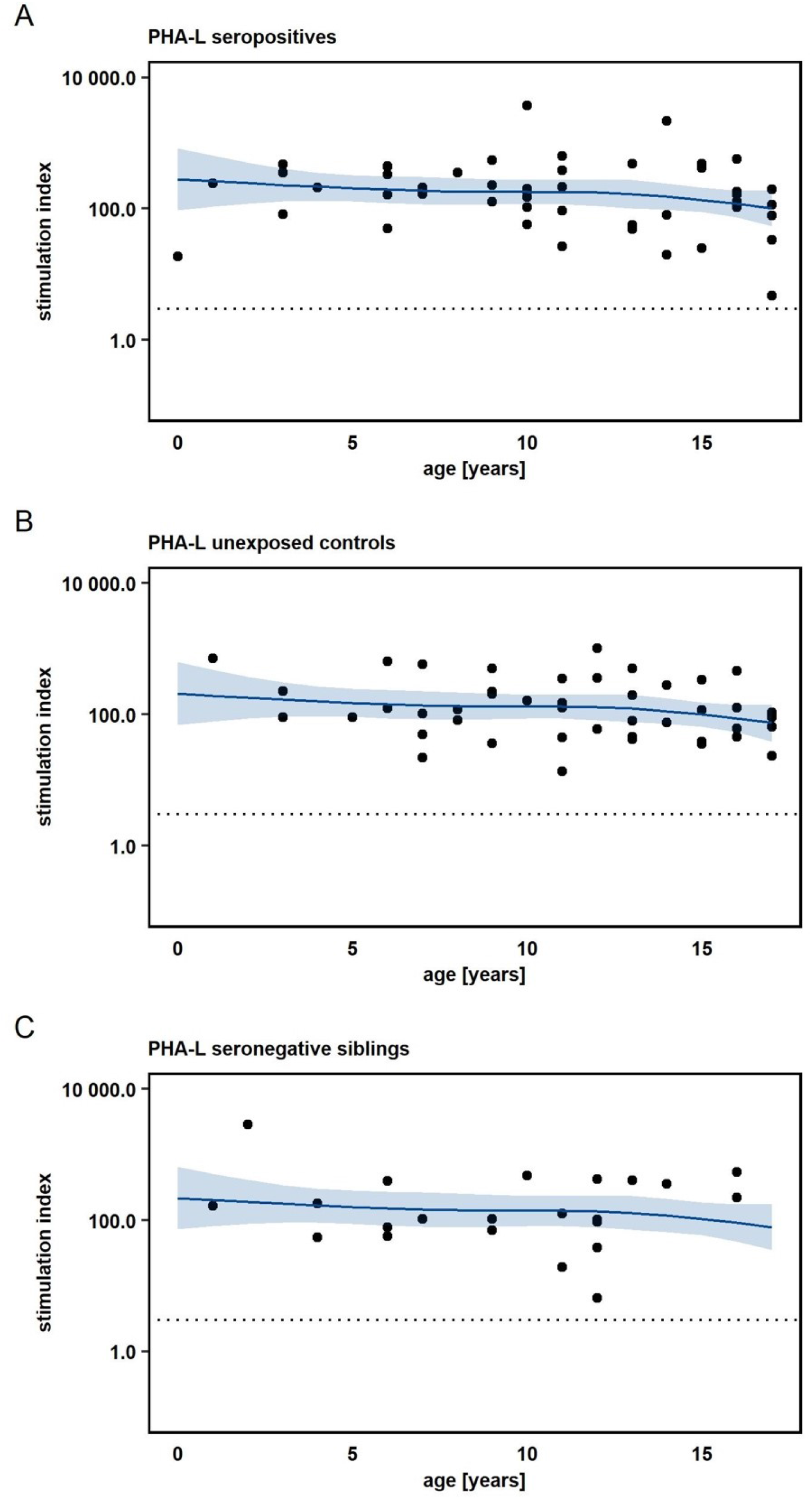
T cell response to stimulation by PHA-L (positive control) according to experimental groups and age Panels according to experimental groups as indicated on top. T cell responses in the positive control, phytohemagglutinin (PHA-L) are displayed as quantified by stimulation index according to the participants’ age. Dots represent individual T cell responses. The effect of age on the magnitude of T cell response was analyzed with a non-parametric multivariate regression analysis, by using a spline model (blue lines with light blue areas indicating 95% confidence intervals).

## References

1 Zhu N, Zhang D, Wang W, Li X, Yang B, Song J et al. A Novel Coronavirus from Patients with Pneumonia in China, 2019. N Engl J Med 2020; 382: 727–733.

2 Johns Hopkins University. COVID-19 Dashboard by the Center for Systems Science and Engineering (CSSE). https://coronavirus.jhu.edu/map.html (Accessed 22 Jan 2022).

3 Preston LE, Chevinsky JR, Kompaniyets L, Lavery AM, Kimball A, Boehmer TK et al. Characteristics and Disease Severity of US Children and Adolescents Diagnosed with COVID-19. JAMA Netw Open 2021; 4: 8–11.

4 Dong Y, Dong Y, Mo X, Hu Y, Qi X, Jiang F et al. Epidemiology of COVID-19 among children in China. Pediatrics 2020; 145. doi:10.1542/peds.2020-0702.

5 CDC Covid-Net. A weekly summary of U.S. COVID-19 hospitalization data; Laboratory-Confirmed COVID-19-Associated Hospitalizations. https://gis.cdc.gov/grasp/COVIDNet/COVID19_3.html (Accessed 22 Jan 2022).

6 Reiss S, Baxter AE, Cirelli KM, Dan JM, Morou A, Daigneault A et al. Comparative analysis of activation induced marker (AIM) assays for sensitive identification of antigen-specific CD4 T cells. PLoS One 2017; 12: 1–22.

7 Braun J, Loyal L, Frentsch M, Wendisch D, Georg P, Kurth F et al. SARS-CoV-2-reactive T cells in healthy donors and patients with COVID-19. Nature 2020. doi:10.1038/s41586-020-2598-9.

8 Grifoni A, Weiskopf D, Ramirez SI, Mateus J, Dan JM, Moderbacher CR et al. Targets of T Cell Responses to SARS-CoV-2 Coronavirus in Humans with COVID-19 Disease and Unexposed Individuals. Cell 2020; 181: 1489–1501.e15.

9 Jung JH, Rha MS, Sa M, Choi HK, Jeon JH, Seok H et al. SARS-CoV-2-specific T cell memory is sustained in COVID-19 convalescent patients for 10 months with successful development of stem cell-like memory T cells. Nat Commun 2021; 12: 1–12.

10 Rydyznski Moderbacher C, Ramirez SI, Dan JM, Grifoni A, Hastie KM, Weiskopf D et al. Antigen-Specific Adaptive Immunity to SARS-CoV-2 in Acute COVID-19 and Associations with Age and Disease Severity. Cell 2020; 183: 996–1012.e19.

11 Schub D, Klemis V, Schneitler S, Mihm J, Lepper PM, Wilkens H et al. High levels of SARS-CoV-2–specific T cells with restricted functionality in severe courses of COVID-19. JCI Insight 2020; 5. doi:10.1172/jci.insight.142167.

12 Steiner S, Sotzny F, Bauer S, Na IK, Schmueck-Henneresse M, Corman VM et al. HCoV- and SARS-CoV-2 Cross-Reactive T Cells in CVID Patients. Front Immunol 2020; 11: 1–10.

13 Thieme CJ, Anft M, Paniskaki K, Blazquez-Navarro A, Doevelaar A, Seibert FS et al. Robust T Cell Response Toward Spike, Membrane, and Nucleocapsid SARS-CoV-2 Proteins Is Not Associated with Recovery in Critical COVID-19 Patients. Cell Reports Med 2020; 1. doi:10.1016/j.xcrm.2020.100092.

14 Weiskopf D, Schmitz KS, Raadsen MP, Grifoni A, Okba NMA, Endeman H et al. Phenotype and kinetics of SARS-CoV-2-specific T cells in COVID-19 patients with acute respiratory distress syndrome. Sci Immunol 2020; 5: eabd2071.

15 Kaaijk P, Olivo Pimentel V, Emmelot ME, Poelen M, Cevirgel A, Schepp RM et al. Children and adults with mild COVID-19 symptoms develop memory T cell immunity to SARS-CoV-2. medRxiv 2021; : 2021.09.10.21263333.

16 Cohen CA, Li APY, Hachim A, Hui DSC, Kwan MYW, Tsang OTY et al. SARS-CoV-2 specific T cell responses are lower in children and increase with age and time after infection. Nat Commun 2021; 12: 1–14.

17 Grifoni A, Sidney J, Zhang Y, Scheuermann RH, Peters B, Sette A. A Sequence Homology and Bioinformatic Approach Can Predict Candidate Targets for Immune Responses to SARS-CoV-2. Cell Host Microbe 2020; 27: 671–680.e2.

18 Sibbertsen F, Glau L, Paul K, Mir TS, Gersting SW, Tolosa E et al. Phenotypic analysis of the pediatric immune response to SARS-CoV-2 by flow cytometry. Cytometry A 2021. doi:10.1002/cyto.a.24528.

19 Bertoletti A, Le Bert N, Qui M, Tan AT. SARS-CoV-2-specific T cells in infection and vaccination. Cell Mol Immunol 2021. doi:10.1038/s41423-021-00743-3.

20 Xia S, Zhang Y, Wang Y, Wang H, Yang Y, Gao GF et al. Safety and immunogenicity of an inactivated SARS-CoV-2 vaccine, BBIBP-CorV: a randomised, double-blind, placebo-controlled, phase 1/2 trial. Lancet Infect Dis 2021; 21: 39–51.

21 Wang H, Zhang Y, Huang B, Deng W, Quan Y, Wang W et al. Development of an Inactivated Vaccine Candidate, BBIBP-CorV, with Potent Protection against SARS-CoV-2. Cell 2020; 182: 713–721.e9.

22 Mulligan MJ, Lyke KE, Kitchin N, Absalon J, Gurtman A, Lockhart S et al. Phase I/II study of COVID-19 RNA vaccine BNT162b1 in adults. Nature 2020; 586: 589–593.

23 Corbett KS, Edwards DK, Leist SR, Abiona OM, Boyoglu-Barnum S, Gillespie RA et al. SARS-CoV-2 mRNA vaccine design enabled by prototype pathogen preparedness. Nature 2020; 586: 567–571.

24 Harvey WT, Carabelli AM, Jackson B, Gupta RK, Thomson EC, Harrison EM et al. SARS-CoV-2 variants, spike mutations and immune escape. Nat Rev Microbiol 2021; 19: 409–424.

25 Rouers A, Chng MHY, Lee B, Rajapakse MP, Kaur K, Toh YX et al. Immune cell phenotypes associated with disease severity and long-term neutralizing antibody titers after natural dengue virus infection. Cell Reports Med 2021; 2. doi:10.1016/j.xcrm.2021.100278.

26 Weng K, Zhang J, Mei X, Wu A, Zhang B, Cai M et al. Lower number of plasmacytoid dendritic cells in peripheral blood of children with bronchiolitis following respiratory syncytial virus infection. Influenza Other Respi Viruses 2014; 8: 469–473.

27 Swain SL, McKinstry KK, Strutt TM. Expanding roles for CD4 + T cells in immunity to viruses. Nat Rev Immunol 2012; 12: 136–148.

28 Dan JM, Mateus J, Kato Y, Hastie KM, Yu ED, Faliti CE et al. Immunological memory to SARS-CoV-2 assessed for up to 8 months after infection. Science (80-) 2021; 371. doi:10.1126/science.abf4063.

29 Le Bert N, Tan AT, Kunasegaran K, Tham CYL, Hafezi M, Chia A et al. SARS-CoV-2-specific T cell immunity in cases of COVID-19 and SARS, and uninfected controls. Nature 2020; 584: 457–462.

30 Jordan SC, Shin BH, Gadsden TAM, Chu M, Petrosyan A, Le CN et al. T cell immune responses to SARS-CoV-2 and variants of concern (Alpha and Delta) in infected and vaccinated individuals. Cell Mol Immunol 2021; 18: 2554–2556.

31 Wang Z, Yang X, Zhong J, Zhou Y, Tang Z, Zhou H et al. Exposure to SARS-CoV-2 generates T-cell memory in the absence of a detectable viral infection. Nat Commun 2021; 12: 6–13.

32 Shannon I, White CL, Murphy A, Qiu X, Treanor JJ, Nayak JL. Differences in the influenza-specific CD4 T cell immunodominance hierarchy and functional potential between children and young adults. Sci. Rep. 2019; 9. doi:10.1038/s41598-018-37167-5.

33 Prabhudas M, Adkins B, Gans H, King C, Levy O, Ramilo O et al. Challenges in infant immunity: Implications for responses to infection and vaccines. Nat Immunol 2011; 12: 189–194.

34 Pichichero ME. Challenges in vaccination of neonates, infants and young children. Vaccine 2014; 32: 3886–3894.

35 Selin LK, Wlodarczyk MF, Kraft AR, Nie S, Kenney LL, Puzone R et al. Heterologous immunity: Immunopathology, autoimmunity and protection during viral infections. Autoimmunity 2011; 44: 328–347.

36 Agrawal B. Heterologous Immunity: Role in Natural and Vaccine-Induced Resistance to Infections. Front Immunol 2019; 10: 1–11.

37 Mateus J, Grifoni A, Tarke A, Sidney J, Ramirez SI, Dan JM et al. Selective and cross-reactive SARS-CoV-2 T cell epitopes in unexposed humans. Science (80-) 2020; 370: 89–94.

38 Loske J, Röhmel J, Lukassen S, Stricker S, Magalhães VG, Liebig J et al. Pre-activated antiviral innate immunity in the upper airways controls early SARS-CoV-2 infection in children. Nat Biotechnol 2021. doi:10.1038/s41587-021-01037-9.

39 Neidleman J, Luo X, McGregor M, Xie G, Murray V, Greene WC et al. mRNA vaccine-induced SARS-CoV-2-specific T cells recognize B.1.1.7 and B.1.351 variants but differ in longevity and homing properties depending on prior infection status. bioRxiv Prepr Serv Biol 2021. doi:10.1101/2021.05.12.443888.

40 Keeton R, Tincho MB, Ngomti A, Baguma R, Suzuki A, Khan K et al. SARS-CoV-2 spike T cell responses induced upon vaccination or infection remain robust against Omicron. 2021; : 1–20.

41 Choi SJ, Kim D, Park S. CORRESPONDENCE T cell epitopes in SARS-CoV-2 proteins are substantially conserved in the Omicron variant. 2022; : 1–2.

42 Greaney AJ, Loes AN, Crawford KHD, Starr TN, Malone KD, Chu HY et al. Comprehensive mapping of mutations in the SARS-CoV-2 receptor-binding domain that affect recognition by polyclonal human plasma antibodies. Cell Host Microbe 2021; 29: 463–476.e6.

43 Scripps Research. outbreak.info - a standardized, open-source database of COVID-19 resources and epidemiology data. https://outbreak.info/situation-reports (Accessed 22 Jan2022).

44 Roarty C, Tonry C, McFetridge L, Mitchell H, Watson C, Waterfield T et al. Kinetics and seroprevalence of SARS-CoV-2 antibodies in children. Lancet Infect. Dis. 2020. doi:10.1016/S1473-3099(20)30884-7.

